# Prevalence and associated factors of mental and substance use problems among adults in Kenya: a community-based cross-sectional study

**DOI:** 10.1101/2024.12.16.24319125

**Authors:** Patrick N. Mwangala, Anita Kerubo, Millicent Makandi, Rachael Odhiambo, Amina Abubakar

## Abstract

**Background:** Data on the burden and determinants of mental and substance use problems among women in urban and rural informal settlements in Kenya is sparse, thus limiting preventive and treatment efforts in these areas. To bridge the gap, we (a) determined the prevalence of depressive, anxiety and post-traumatic stress disorder (PTSD) symptoms and alcohol and drug use problems among women compared to their spouses and (b) examined the risk and protective indicators associated with these outcomes.

**Methods:** Data collection for this cross-sectional survey was conducted in 2022 in Mombasa, Kwale and Nairobi counties in Kenya. A total of 1528 adults (1048 women) took part. The 9-Item Patient Health Questionnaire, 7-Item Generalized Anxiety Disorder Scale, Primary Care PTSD Screen for DSM-5, Alcohol Use Disorders Identification Test, and Drug Use Disorders Identification Test were administered alongside other measures. Logistic regression was used to examine the correlates of mental and substance use problems.

**Results:** Overall, the prevalence of mental and substance use problems was 28% vs 22% (depressive symptoms), 12% vs 8% (general anxiety symptoms), 22% vs 21% (PTSD symptoms), 4% vs 15% (alcohol use problems), and 2% vs 12% (drug use problems) among women and their spouses respectively. The prevalence of depressive and anxiety symptoms was significantly higher among women compared to their male counterparts. On the other hand, both current and past-year alcohol and drug use were significantly higher in men than women. Among women, stressful life events, urban residence, food insecurity, family debt, unemployment, poor self-rated health, poor eyesight, and higher educational level were the risk indicators for elevated depressive, anxiety and PTSD symptoms. Conversely, sexual abuse, living in rented houses, urban residence, verbal abuse, stressful life events, and somatic complaints were the risk indicators for depressive, anxiety and PTSD symptoms in men. Protective indicators against mental health problems included social support, higher subjective wellbeing, older age (>50 years), increased vigorous exercise and higher household income (in both sexes). Risk indicators for current alcohol use in women included stressful life events, urban residence, being sexually active, and living in a single family. Among men, higher household income was associated with current alcohol use. Protective indicators against current alcohol use included being married, living in a larger household (>5), being a Muslim and having multimorbidity (in both sexes). Risk indicators for current drug use included unemployment and sexual abuse. Female sex was associated with reduced odds of current drug use.

**Conclusion:** The burden of mental and substance use problems in women and their spouses is elevated for most of the conditions assessed. However, needs vary by gender and study location, highlighting the importance of targeted approaches in mental health services. Our results also highlight the need for multicomponent preventive and treatment strategies to mitigate the risks of mental and substance use problems in this population.

## Background

The importance of mental health as an integral part of general health and sustainable development has long been recognized [1–3]. In itself, mental health is essential for physical health and is closely connected with development factors such as work, economic growth, poverty, peace and justice. Mental health also plays a crucial role in achieving social inclusion, equity, universal health coverage, access to justice and human rights. Indeed, the mental health landscape has transformed over the last three decades thanks to higher visibility of mental health problems, political goodwill and multilateral commitments. As substantial as these efforts are, several gaps and challenges still exist that warrant more revitalized actions from stakeholders. For example, since the first Global Burden of Disease Study findings were published in the 1990s, there has been increasing evidence that mental and substance use disorders constitute a leading cause of disease burden around the world and about three-quarters of this burden lies in low-and-middle-income countries (LMICs) [4, 5]. This burden is high across the lifespan, for both males and females and across many societies. Importantly, there is no evidence of a reduction in the global prevalence or burden of these disorders since 1990, despite compelling evidence of effective interventions [6–8]. Furthermore, the burden of mental and substance use disorders is projected to rise globally in the coming decades, and the largest increase can be expected in LMICs as a result of population growth, rising life expectancy and under-resourced healthcare systems [9]. Predictive models indicate a 130% rise in the burden of mental and substance disorders in sub-Saharan Africa (SSA) by 2050 because of population growth and ageing [10]. At the same time, there is a large treatment gap for these disorders – it is estimated that only 1% of the global health workforce provides mental healthcare, a contributing factor to the estimated 75% to 95% treatment gap [11–13]. Where treatments are accessed, sometimes they lack a clear evidence base and involve significant out-of-pocket payments, which can cause catastrophic health expenditure, especially in LMICs [14].

The Eastern SSA region is undergoing a rapid epidemiological transition from communicable to non-communicable disease burden [15]. Kenya is one such country that is witnessing this transition – with improving life expectancy, reduced malnutrition, and a concomitant rise in the burden of non-communicable diseases [16]. According to the most recent census of 2019, Kenya had a population of 47.5 million people, nearly 60% of whom were 24 years old or younger [17]. In recent years, the conversation around mental and substance use disorders has increasingly taken centre stage, given the realization that mental health issues are pervasive and impactful. For instance, data from the Kenya National Commission on Human Rights estimates that 25% and 40% of outpatients and inpatients suffer from mental health problems, respectively [18]. The frequently reported mental health problems include depression, anxiety, and substance abuse. A recent WHO report ranked Kenya fifth among African countries with increased cases of depression [19]. The Kenya Adolescent Mental Health Group recently analyzed the burden and risk factors of mental and substance use disorders among adolescents and young adults from the Global Burden of Disease Study of 2019 [20]. The authors found that mental disorders ranked as the second leading cause of disability in this population after unintentional injuries. A recent review also reported a significant burden of substance use among adults and adolescents in Kenya [21]. In response, the government of Kenya has progressively made efforts to improve mental health services in the country. These efforts include the launch of several policy and legislative frameworks, including the Kenya Mental Health Policy 2015-2030, Kenya Mental Health Action Plan 2021-2025, Kenya Mental Health Investment Case 2021, Mental Health (Amendment) Act of 2022, Suicide Prevention Strategy 2021-2026, the 2018 National Framework for Implementation of Problem management Plus and the 2019 presidential taskforce on Mental Health in Kenya [18]. While this is laudable, like other SSA countries, Kenya still struggles with the potent implementation of the existing mental health policies, leading to a continually widening mental health treatment gap [22, 23]. Currently, about 75% of Kenyans are not able to access mental healthcare, nearly half of the counties do not have psychiatric units, and the country still does not have a separate mental health budget, and government expenditure on mental health is 0.01% of the health expenditure.

Generally, the available data on the burden and determinants of mental and substance use disorders in Kenya is sparse. For example, the national-level survey of non-communicable diseases in Kenya did not include mental health [24]. Epidemiological research is crucial to understand better the differential risk factors and burden of mental and substance use disorders across different regions and social contexts. Data on adolescent mental health is building up, thanks to the establishment of nationally representative surveys [25]. While this is encouraging, more data is needed to generate more accurate estimates of health burdens and salient determinants from which local health policymakers can draw for other populations. Such research will inform optimal strategies for prevention, treatment and follow-up for people with mental and substance use disorders across the life course. Most of the existing studies have been collected from small samples of diverse sets of populations, including people living with HIV, youth, and pregnant women, although not necessarily representative of the population. There is especially a dearth of research on the burden and determinants of mental and substance use disorders among adults in the general population. The current study addresses this gap by a) documenting the prevalence of mental health problems (symptoms of depression, anxiety and post-traumatic stress disorder) and substance use problems (alcohol and drug use problems) in a community sample of adults from Nairobi, Mombasa and Kwale counties in Kenya, and b) investigate the factors associated with these problems.

## Methods

### Study design and setting

This was a cross-sectional study conducted in 2022 in Nairobi, Mombasa and Kwale counties in Kenya. The data used in this study was obtained from the formative phase of the ’Advancing Gender Equality through Civil Society’ (AGECS) mental health research project being implemented at the Aga Khan University, Institute for Human Development in Kenya. AGECS is a mixed methods research project that seeks to understand women’s mental health needs and tailor programmes that address these needs. Further details of the project have been described elsewhere [26]. Nairobi and Mombasa counties are predominantly urban, while Kwale is largely rural. In Nairobi, the study was conducted in Westlands, one of the county’s seventeen sub-counties. In Mombasa, the study was carried out in Changamwe, one of the county’s six sub-counties. In Kwale, the study was conducted in Matuga, one of the county’s five sub-counties.

### Sample size

We estimated the sample size using power analyses in Stata based on previous studies. An overall sample of at least 1000 participants was required to detect a difference in mental and substance use problems between males and females at 80% power and a 5% level of statistical significance. A final sample of 1528 was considered sufficient, allowing for missing data and conducting stable psychometric models for some of the measures.

### Participants recruitment and data collection procedures

Our target population in the cross-sectional survey was adult men and women aged 18 years and older living in the three counties of interest. Potential clients were recruited using sequential sampling from households in the three counties. Participant recruitment went on until the desired sample size was attained. Recruitment of participants was carried out by community health volunteers (CHVs) at the household level. We drew on the CHVs’ knowledge of both the geography and local population of the study sites to identify potential clients. To be included in the survey, participants had to be at least 18 years old, be able to provide informed consent, and be able to speak Swahili or English. The majority of the assessments were taken in Swahili. Participants recruitment began on 27^th^ September 2022 and the last person was recruited on 8^th^ December 2022.

Once recruited by CHVs, the participants were booked for assessment, usually the following day, at a central venue in the community, which included social halls. All participants who turned up on the day of the assessment consented to the study by a team of 11 research assistants. The team of research assistants received a 1-week training in both sites, Nairobi and the Coast of Kenya (Mombasa and Kwale). Topics covered during the training sessions included an overview of psychological constructs, a review of measures, psychological first aid, research ethics, data management and how to conduct face-to-face interviews. They had a chance to practice this through role plays. Pre-testing of tools was also done through a pilot study before the eventual implementation of the study.

All the data was collected face-to-face utilizing the Open Data Kit through tablets that were password-protected and encrypted to avoid data loss. The assessments took approximately an hour. Thereafter, a small refreshment was given to the participants. Participants were also reimbursed for their transport costs. A data manager double-checked any inconsistencies in the data before uploading it to the server on a daily basis.

The final sample included in the analysis comprised of 1048 women and 480 men. 353 were from Kwale County, 583 from Mombasa County and 592 from Nairobi County. Further recruitment details are captured in Figure 1.

**Figure 1.**
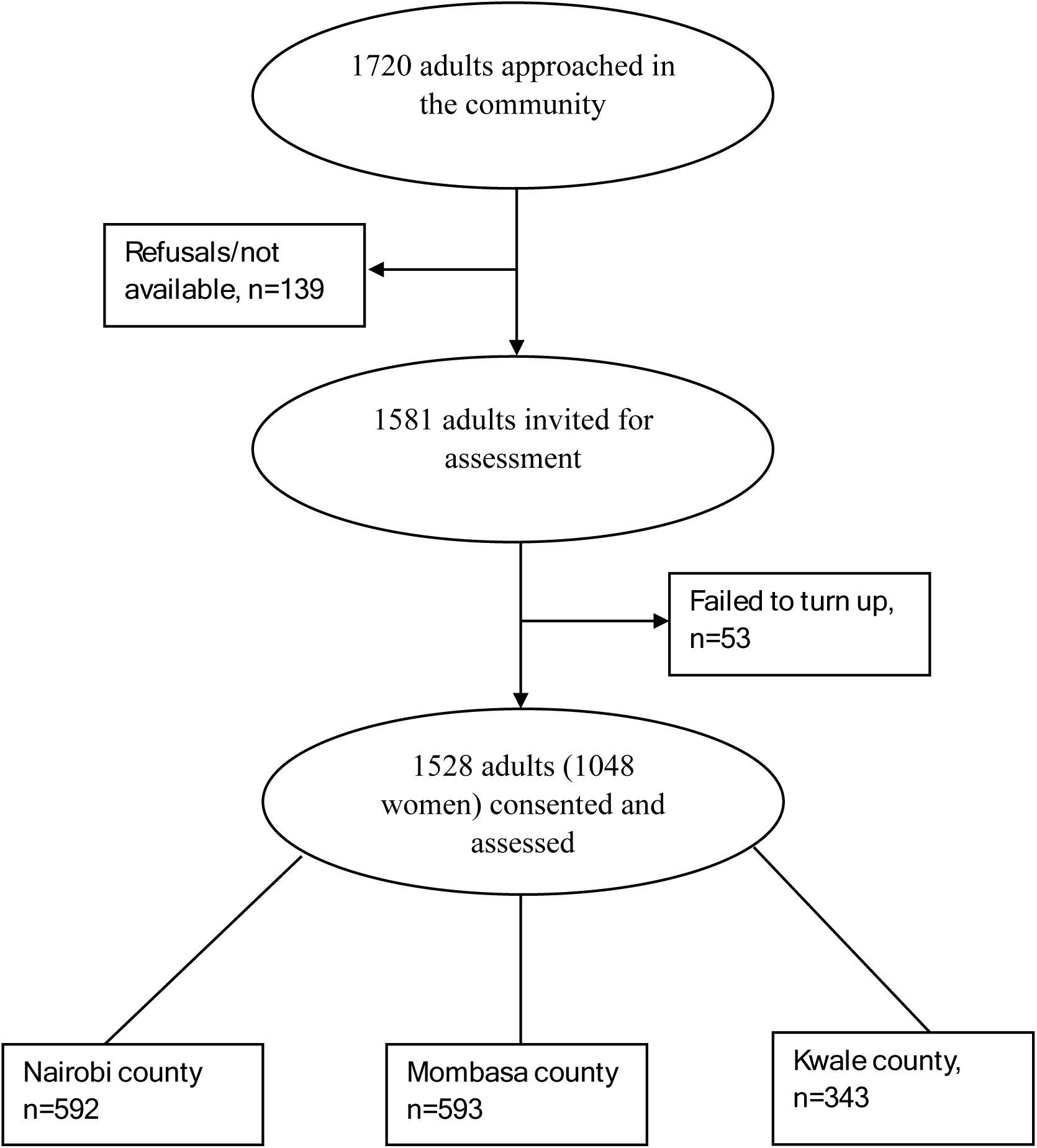
Participant recruitment flowchart

## Measures

### Sociodemographic characteristics

We collected data on the participant’s age, sex, marital status, educational level, religion, participant’s occupation, household size, household income, type of family, living arrangements, family debt, and caregiving status. Participants’ socioeconomic status was assessed using an asset index that has been previously used in the study settings. The tool screened for the ownership of a list of disposable assets by participants (or their families) such as radio, television, bicycle, and motorbike. A single score of socioeconomic status was generated, with a higher score translating to a higher socioeconomic status.

### Lifestyle and health history information

Information collected included participant’s self-reported health status, lifetime experience of abuse (verbal, physical, sexual), self-reported hearing and vision, history of any chronic condition (e.g. diabetes, hypertension, cancer, HIV), somatic complaints (e.g. migraine, fatigue, pain), and self-reported physical activity. Additional information sought from women included pregnancy status, age at first pregnancy, pregnancy in the previous year, number of live pregnancies, sexual activity, and history of infertility.

## Outcome measures

### Depressive symptoms

We used the 9-item patient health questionnaire (PHQ-9) to screen for depressive symptoms among participants. The tool has 9 items that are scored on a 4-point Likert scale (0 =not at all, 3 = nearly every day). The total scores range from 0 to 27. Total scores of 5-9, 10-14, and 15-27 indicate mild, moderate and severe depressive symptoms, respectively. The Swahili version of PHQ-9 has been validated extensively in Kenya, and there is empirical evidence that it is suitable for use among different populations, including adolescents [27], adults [28] and healthcare providers [29]. In the current study, the tool registered good internal consistency (Cronbach’s alpha = 0.83).

### Anxiety symptoms

The 7-item generalized anxiety disorder questionnaire (GAD-7) was used to screen for anxiety among respondents. The items are scored on a 4-point Likert scale ranging from 0 (not at all) to 3 (more than half the days). The total scores range from 0 to 21, with cut-offs of 5-9, 10-14 and 15-21 representing mild, moderate and severe anxiety symptoms, respectively. The tool has previously been validated in Kenya, maintaining its unidimensional latent structure [30]. In the present study, the tool demonstrated good internal reliability with an alpha value of 0.87.

### Symptoms of post-traumatic stress disorder (PTSD)

We used the 5-item primary care PTSD screen (PC-PTSD-5) to assess for PTSD. Participants first complete a screening question to determine lifetime exposure to trauma. Respondents who have not experienced any exposure to trauma are assigned a score of zero. Those who acknowledge exposure to trauma in the past month are asked five questions, which are scored dichotomously (0=no; 1=yes). The total scores range from 0 to 5. We used a cut-off of ≥3 to indicate a positive screen for PTSD in the current study, similar to previously published studies [31–33]. PC-PTSD-5 was validated in the current study, yielding good psychometric properties [26].

### Alcohol use problems

The 10-item Alcohol Use Disorders Identification Test (AUDIT) was used to screen for patterns of hazardous/harmful alcohol use and alcohol dependence. All participants first responded to a screening question (yes/no) assessing whether they currently used any alcoholic drink. Participants responding yes went ahead and answered the subsequent AUDIT items. The tool has a total score of 40. A total score of ≥8 and ≥6 for males and females, respectively, was used to define a positive screen for hazardous drinking, similar to previously published studies [34].

### Drug use problems

We used the 11-item Drug Use Disorders Identification Test (DUDIT) to screen for hazardous drug use. Like the AUDIT, items are summated with a maximum score of 44. We used a cut-off score of ≥6 and ≥2 for males and females, respectively, to define a probable positive screen for problematic drug use, similar to previous research [34].

## Psychosocial measures

*The 17-item Intimate Partner Violence Attitude Scale – Revised (IPVAS—R)* was used to assess participants’ attitudes towards intimate partner violence [35]. Higher scores indicate a greater level of negative attitudes towards IPV.

The *12-item Multidimensional Scale of Perceived Social Support (MSPSS)* [36] was used to assess participants’ social support. A higher score indicates greater support perceived by an individual.

*The 5-item WHO Wellbeing Index* [37] was used to assess participants’ subjective wellbeing. A higher score indicates greater wellbeing by a participant.

*The 13-item Stressful Life Events Screening Questionnaire (SLESQ*) [38] was used to assess traumatic event exposure among participants. A summated score was generated to show the total number of life events reported.

## Ethics

The study was approved by the Aga Khan, Nairobi Institutional Scientific and Ethics Review Committee (Ref: 2022/ISERC_44(V2)). Permission to carry out the study in Kenya was sought and granted by the National Commission for Science, Technology and Innovation (Ref: 346643). Respective County departments of health gave local permits to conduct the study: Nairobi (Ref: NCCG/DHS/REC/240), Mombasa (Ref: MCG/COPH/RCH./111) and Kwale (Ref: CG/KWL/6/6/1/CECM/39/VOL.1/34). All respondents provided written informed consent for their participation in the study.

## Data analysis

We analyzed our data using STATA version 17.0 (StataCorp LP, College Station, TX, United States). Sociodemographic and health history variables were summarized using descriptive statistics: mean and standard deviation for continuous variables and frequency and proportion for categorical variables. We used proportions to estimate the prevalence of mental and substance use problems among participants. We used logistic regression models to explore univariate associations between binary outcome variables and various exposure variables. Independent variables with a p-value <0.15 in the univariate analysis were then fitted into the multivariable models using forward selection. Collinearity was checked in all models, and a two-tailed p-value of <0.05 was deemed statistically significant for all hypothesis tests. The overall fit of the final models was assessed using Hosmer and Lemeshow’s goodness of fit, and a p-value of >0.05 was considered a good fit.

## Results

### Sample characteristics

The characteristics of the included participants are presented in **Table 1**. The overall sample comprised 1528 participants (68.6% women), with a mean age of 39.1 years (SD = 13.1). The vast majority of the participants were from urban informal settlements (77.6%), had attained basic education or more (94.7%), were married (75.4%), Christians (64.1%), living in nuclear families (74.8%), living in rented houses (67.8%), in debt (83.1%) and experiencing food insecurity (71.7%). Women were likely to be unemployed, have lower monthly household income, have lower socioeconomic status, report poor self-rated health status, experience physical abuse from a spouse, and have chronic conditions. Further details are highlighted in Table 1.

**Table 1.** Characteristics of the study population by gender, *n* = 1528.

| Characteristic | Total sample<br>*n* = 1528 | Gender |  |  |
| --- | --- | --- | --- | --- |
|  |  | *Females, n* = 1048 | *Males, n* = 480 | *p-value* |
| Sociodemographic |  |  |  |  |
| Area of residence |  |  |  |  |
| *Kwale (rural)* | 343 (22.4%) | 254 (24.2%) | 89 (18.5%) | 0.01 |
| *Mombasa & Nairobi (Urban)* | 1185 (77.6%) | 794 (75.8%) | 391 (81.5%) |  |
| Age in years, missing = 2 |  |  |  |  |
| *18 – 34* | 657 (43.1%) | 484 (46.3%) | 173 (36.0%) | <0.001 |
| *35 – 49* | 536 (35.1%) | 359 (34.3%) | 177 (36.9%) |  |
| *≥50* | 333 (21.8%) | 203 (19.4%) | 130 (27.1%) |  |
| Level of education |  |  |  |  |
| *Tertiary* | 196 (12.8%) | 130 (12.4%) | 66 (13.8%) | 0.001 |
| *Secondary* | 521 (34.1%) | 328 (31.3%) | 193 (40.2%) |  |
| *Primary* | 731 (47.8%) | 525 (50.1%) | 206 (42.9%) |  |
| *None* | 80 (5.3%) | 65 (6.2%) | 15 (3.1%) |  |
| Employment |  |  |  |  |
| *Skilled/Professional* | 315 (20.6%) | 165 (15.7%) | 150 (31.3%) | <0.001 |
| *Unskilled/casual* | 705 (46.1%) | 457 (43.6%) | 248 (51.7%) |  |
| *Unemployed* | 508 (33.3%) | 426 (40.75) | 82 (17.0%) |  |
| Marital status, missing = 5 |  |  |  |  |
| *Never married* | 103 (6.8%) | 97 (9.3%) | 6 (1.3%) | <0.001 |
| *Separated/Divorced/Widowed* | 271 (17.8%) | 269 (25.7%) | 2 (0.4%) |  |
| *Married/cohabiting* | 1149 (75.4%) | 680 (65.0%) | 469 (98.3%) |  |
| Religion |  |  |  |  |
| *Christian* | 980 (64.1%) | 670 (63.9%) | 310 (64.6%) | 0.37<sup>§</sup> |
| *Muslim* | 545 (35.7%) | 377 (36.0%) | 168 (35.0%) |  |
| *No religion* | 3 (0.2%) | 1 (0.1%) | 2 (0.4%) |  |

| Table 1. Characteristics of the study population by gender, n = 1528 (Continued) |  |  |  |  |
| --- | --- | --- | --- | --- |
| Characteristic | Total sample<br>n = 1528 | Gender |  | p-value |
|  |  | Females, n = 1048 | Males, n = 480 |  |
| Type of family |  |  |  |  |
| Nuclear | 1143 (74.8%) | 722 (68.9%) | 421 (87.7%) | <0.001 |
| Extended | 245 (16.0%) | 187 (17.8%) | 58 (12.1%) |  |
| Single | 140 (9.2%) | 139 (13.3%) | 1 (0.2%) |  |
| Living arrangements |  |  |  |  |
| Own house (not paying rent) | 492 (32.2%) | 344 (32.8%) | 148 (30.8%) | 0.44 |
| In a rented house | 1036 (67.8%) | 704 (67.2%) | 332 (69.2%) |  |
| Size of household |  |  |  |  |
| 1 – 2 | 193 (12.6%) | 143 (13.7%) | 50 (10.4%) | 0.19 |
| 3 – 4 | 640 (41.9%) | 430 (41.0%) | 210 (43.8%) |  |
| ≥5 | 695 (45.5%) | 475 (45.3%) | 220 (45.8%) |  |
| Monthly household income (Ksh) |  |  |  |  |
| <10000 | 976 (63.9%) | 727 (69.4%) | 249 (51.9%) | <0.001 |
| 10000 – 29999 | 490 (32.1%) | 281 (26.8%) | 209 (43.5%) |  |
| >30000 | 62 (4.0%) | 40 (3.8%) | 22 (4.6%) |  |
| Household in debt, missing =20 |  |  |  |  |
| Yes | 1253 (83.1%) | 848 (82.3%) | 405 (84.7%) | 0.25 |
| No | 255 (16.9%) | 182 (17.7%) | 73 (15.3%) |  |
| Household food insecurity<br>(last 3 months) |  |  |  |  |
| Yes | 1095 (71.7%) | 759 (72.4%) | 336 (70%) | 0.33 |
| No | 433 (28.3%) | 289 (27.6%) | 144 (30%) |  |
| Family caring for a child living<br>with a disability or chronic<br>illness |  |  |  |  |
| Yes | 88 (5.8%) | 71 (6.8%) | 17 (3.5%) | 0.01 |
| No | 1440 (94.2%) | 977 (93.2%) | 463 (96.5%) |  |
| Asset index score – mean (SD) | 2.9 (1.4) | 2.8 (1.3) | 3.2 (1.4) | <0.001 |
| Health history |  |  |  |  |
| Self-reported health status |  |  |  |  |
| Good | 411 (26.9%) | 264 (25.2%) | 147 (30.6%) | 0.01 |
| Average | 968 (63.3%) | 667 (63.7%) | 301 (62.7%) |  |
| Poor | 149 (9.8) | 117 (11.1%) | 32 (6.7%) |  |
| Lifetime experience of verbal<br>abuse from a spouse |  |  |  |  |
| Yes | 830 (54.3%) | 550 (52.5%) | 280 (58.3%) | 0.03 |
| No | 698 (45.7%) | 498 (47.5%) | 200 (41.7%) |  |
| Lifetime experience of physical<br>abuse from a spouse |  |  |  |  |
| Yes | 430 (28.1%) | 385 (36.7%) | 45 (9.4%) | <0.001 |
| No | 1098 (71.9%) | 663 (63.3%) | 435 (90.6%) |  |
| Lifetime experience of sexual<br>abuse from a spouse |  |  |  |  |
| Yes | 216 (14.1%) | 140 (13.4%) | 76 (15.8%) | 0.20 |
| No | 1312 (85.9%) | 908 (86.6%) | 404 (84.2%) |  |

| Table 1. Characteristics of the study population by gender, n = 1528 (*Continued*) |  |  |  |  |
| --- | --- | --- | --- | --- |
| Characteristic | Total sample<br>*n* = 1528 | Gender |  |  |
|  |  | *Females, n* = 1048 | *Males, n* = 480 | *p-value* |
| Self-reported vision |  |  |  |  |
| *Good* | 867 (56.7%) | 567 (54.1%) | 300 (62.5%) | **0.01** |
| *Average* | 561 (36.7%) | 406 (38.7%) | 155 (32.3%) |  |
| *Poor* | 100 (6.6%) | 75 (7.2%) | 25 (5.2%) |  |
| Self-reported hearing |  |  |  |  |
| *Good* | 1347 (88.1%) | 920 (87.8%) | 427 (89.0%) | 0.57<sup>§</sup> |
| *Average* | 157 (10.3%) | 109 (10.4%) | 48 (10.0%) |  |
| *Poor* | 24 (1.6%) | 19 (1.8%) | 5 (1.0%) |  |
| Participant living with a chronic condition |  |  |  |  |
| *Yes* | 549 (35.9%) | 405 (38.7%) | 144 (30%) | **0.001** |
| *No* | 979 (64.1%) | 643 (61.3%) | 336 (70%) |  |
| Presence of somatic complaints |  |  |  |  |
| *Yes* | 812 (53.1%) | 593 (56.6%) | 219 (45.6%) | **<0.001** |
| *No* | 716 (46.9%) | 455 (43.4%) | 261 (54.4%) |  |
| Average number of days engaged in vigorous intensity physical activity (in the past 7 days), *Mean (SD)* | 1.5 (2.3) | 1.2 (2.1) | 2.3 (2.6) | **<0.001** |
| Average number of days engaged in moderate intensity physical activity (in the past 7 days), *Mean (SD)* | 3.7 (2.6) | 3.9 (2.5) | 3.2 (2.8) | **<0.001** |
| Average number of days engaged in light intensity physical activity (in the past 7 days), *Mean (SD)* | 4.9 (2.5) | 4.7 (2.6) | 5.4 (2.3) | **<0.001** |
| Reproductive information for females |  |  |  |  |
| Ever pregnant before |  |  |  |  |
| *Yes* | — | 984 (93.9%) | — | N/A |
| *No* | — | 64 (6.1%) | — |  |
| Age at first pregnancy |  |  |  |  |
| *<18 years* | — | 407 (41.4%) | — | N/A |
| *Above 18 years* | — | 577 (58.6%) | — |  |
| Pregnant in the previous year |  |  |  |  |
| *Yes* | — | 174 (17.7%) | — | N/A |
| *No* | — | 810 (82.3%) | — |  |
| Difficulty conceiving for the past 1 year |  |  |  |  |
| *Yes* | — | 71 (6.8%) | — | N/A |
| *No* | — | 977 (93.2%) | — |  |
| Currently sexually active |  |  |  |  |
| *Yes* | — | 695 (66.3%) | — | N/A |
| *No* | — | 353 (33.7%) | — |  |

| **Table 1. Characteristics of the study population by gender, n = 1528 (Continued)** |  |  |  |  |
| --- | --- | --- | --- | --- |
| **Characteristic** | **Total sample**<br>**n = 1528** | **Gender** |  |  |
|  |  | *Females, n = 1048* | *Males, n = 480* | *p-value* |
| **WHO-5 Wellbeing Index – Mean (SD)** | 12.3 (6.5) | 12.0 (6.5) | 12.9 (6.5) | **0.01** |
| **Multidimensional Scale of Perceived Social Support – mean (SD)** | 61.0 (12.0) | 60.0 (12.3) | 63.2 (10.8) | **<0.001** |
| **Stressful life events scale – mean (SD)** | 3.0 (2.4) | 2.9 (2.4) | 3.4 (2.4) | **<0.001** |
| **Intimate Partner Violence Attitude Scale – Mean (SD)** | 36.0 (6.8) | 35.9 (6.8) | 36.1 (6.9) | 0.61 |
| Note: *P*-values are for differences between women and men. The *P*-values have been derived from the Chi-square test (of Fisher's exact test) and independent Student's t-test for categorical and continuous variables, respectively. Abbreviations: Ksh, Kenya shillings; SD, standard deviation. |  |  |  |  |
| § Based on Fisher's exact test. |  |  |  |  |

### Prevalence of mental and substance use problems

**Table 2** presents the prevalence estimates for mental, and substance use problems across the sample and disaggregated estimates by gender (women vs. men). Group differences in mental and substance use problems are also compared statistically and presented in this table.

**Table 2:**
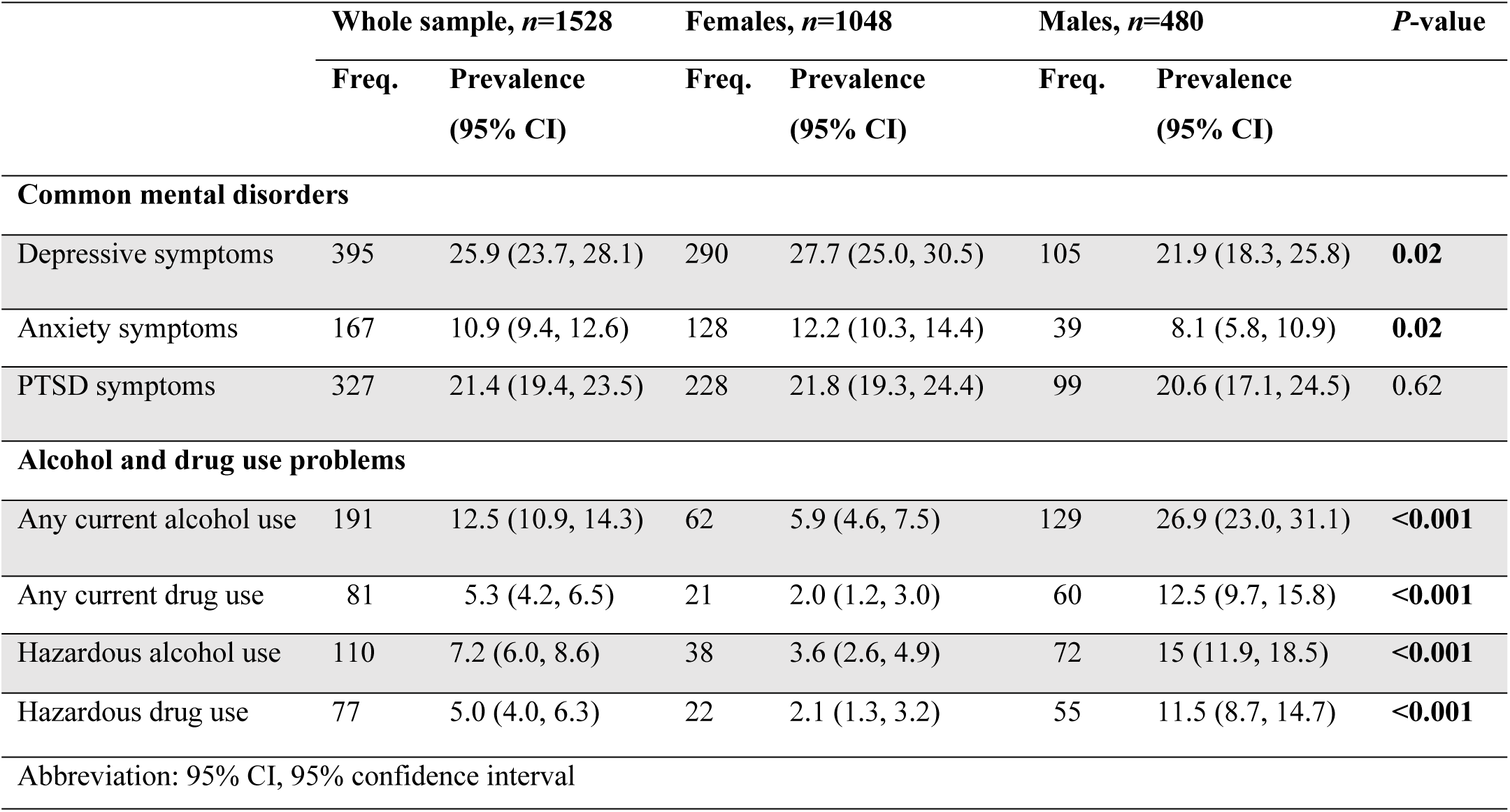
Prevalence of mental and substance use problems among participants.

### Mental health problems

The overall prevalence of depressive symptoms was *25.9% (95% CI 23.7, 28.1%)*, and that for anxiety symptoms was *10.9% (95% CI 9.4, 12.6%).* The overall prevalence of symptoms of PTSD was *21.4% (95% CI 19.4, 23.5%)*. Notably, women reported significantly higher frequency of depressive and anxiety symptoms than men (Table 2).

### Substance use problems

The overall point prevalence of any current alcohol use was *12.5% (95% CI 10.9, 14.3%)* and that for any current drug use was *5.3% (95% CI 4.2, 6.5%)*. Men reported significantly higher prevalence of any current alcohol use and drug use than women (Table 2). The overall past-year prevalence of hazardous alcohol and drug use was 7.2% (95% CI 6.0, 8.6%) and 5.0% (95% CI 4.0, 6.3%), respectively. Similarly, men reported significantly higher frequency of past-year hazardous alcohol and drug use than women (Table 2).

### Risk and protective indicators for mental and substance use problems

**Table 3** summarizes the correlates of mental health problems among women, while **Table 4** summarizes the results for men. **Table 5** presents the correlates of any current substance in both men and women.

**Table 3.** Univariate and multivariable analysis of correlates of mental health problems among women, *n=1048*.

| Covariate | Positive screen for depressive symptoms |  | Positive screen for anxiety symptoms |  | Positive screen for PTSD symptoms |  |
| --- | --- | --- | --- | --- | --- | --- |
|  | Univariate analysis OR (95% CI) | Multivariable analysis aOR (95% CI) | Univariate analysis OR (95% CI) | Multivariable analysis aOR (95% CI) | Univariate analysis OR (95% CI) | Multivariable analysis aOR (95% CI) |
| **Residence** |  |  |  |  |  |  |
| *Rural setting* | Ref | Ref | Ref | Ref | Ref | Ref |
| *Urban setting* | 3.64 (2.42, 5.47)** | 1.70* (1.14, 2.53)<br>3.35** (2.08, 5.39) | 3.14** (1.74, 5.68) | 2.76* (1.41, 5.40) | 2.18** (1.46, 3.25) | 1.72* (1.11, 2.65) |
| **Age in years** |  |  |  |  |  |  |
| *18 – 34* | Ref | Ref | – | – | Ref | Ref |
| *35 – 49* | 1.91** (1.41, 2.58) | 1.24 (0.85, 1.81) | – | – | 1.27 (0.92, 1.76) | 0.89 (0.63, 1.28) |
| *≥50* | 1.11 (0.76, 1.63) | 0.71 (0.42, 1.23) | – | – | 0.72 (0.47, 1.11) | 0.55* (0.33, 0.90) |
| **Educational level** |  |  |  |  |  |  |
| *None* | Ref | Ref | Ref | Ref | Ref | Ref |
| *Primary* | 3.97* (1.68, 9.39) | 3.46* (1.32, 9.10) | 9.20* (1.26, 67.46) | 8.79* (1.16, 66.79) | 4.67* (1.66, 13.09) | 3.58* (1.22, 10.54) |
| *Secondary* | 4.31** (1.80, 10.32) | 4.21* (1.55, 11.45) | 9.66* (1.31, 71.43) | 11.11* (1.43, 86.30) | 4.92* (1.73, 13.95) | 3.33* (1.10, 10.05) |
| *Tertiary* | 3.35* (1.32, 8.46) | 3.54* (1.20, 10.46) | 10.29* (1.34, 78.86) | 15.15* (1.85, 123.97) | 2.94 (0.96, 8.95) | 2.06 (0.63, 6.72) |
| **Occupation** |  |  |  |  |  |  |
| *Skilled/Professional* | – | – | Ref | Ref | – | – |
| *Unskilled/casual labourer* | – | – | 2.04* (1.10, 3.81) | 2.02* (1.02, 4.01) | – | – |
| *Unemployed* | – | – | 1.45 (0.76, 2.76) | 2.13* (1.05, 4.33) | – | – |
| **Type of family** |  |  |  |  |  |  |
| *Nuclear* | – | – | – | – | Ref | Ref |
| *Extended* | – | – | – | – | 0.65 (0.42, 1.01) | 0.76 (0.47, 1.22) |
| *Single* | – | – | – | – | 1.82* (1.23, 2.71) | 1.52* (1.00, 2.32) |
| **Self-reported vision** |  |  |  |  |  |  |
| *Good* | Ref | Ref | – | – | – | – |
| *Average* | 1.07 (0.80, 1.43) | 0.82 (0.57, 1.19) | – | – | – | – |
| *Poor* | 4.06** (2.47, 6.66) | 2.03* (1.07, 3.83) | – | – | – | – |
| **Stressful life events** |  |  |  |  |  |  |
| *None* | Ref | Ref | Ref | Ref | Ref | Ref |
| *>5 events* | 1.62 (0.98, 2.66) | 1.14 (0.65, 1.97) | 1.96 (0.87, 4.44) | 1.42 (0.61, 3.32) | 2.78* (1.45, 5.34) | 2.65* (1.36, 5.14) |
| *≥5 events* | 4.83** (2.94, 7.95) | 2.42* (1.38, 4.23) | 5.86** (2.63, 13.03) | 3.04* (1.31, 7.04) | 7.16** (3.73, 13.71) | 5.95** (3.06, 11.60) |
| **Self-perceived wellbeing** | 0.87** (0.85, 0.89) | 0.90** (0.87, 0.92) | 0.84** (0.81, 0.88) | 0.87** (0.83, 0.91) | – | – |
| **Sexually active** |  |  |  |  |  |  |
| *Yes* | Ref | Ref | – | – | – | – |
| *No* | 1.73** (1.30, 2.28) | 1.40 (0.99, 1.99) | – | – | – | – |

**Table 3 continued**
| Covariate | Positive screen for depressive symptoms |  | Positive screen for anxiety symptoms |  | Positive screen for PTSD |  |
| --- | --- | --- | --- | --- | --- | --- |
|  | Univariate analysis OR (95% CI) | Multivariable analysis aOR (95% CI) | Univariate analysis OR (95% CI) | Multivariable analysis aOR (95% CI) | Univariate analysis OR (95% CI) | Multivariable analysis aOR (95% CI) |
| **Self-rated health status** |  |  |  |  |  |  |
| *Good* | Ref | Ref | Ref | Ref | Ref | Ref |
| *Average* | 2.33** (1.59, 3.42) | 1.74* (1.12, 2.71) | 2.05* (1.17, 3.59) | 1.33 (0.73, 2.43) | 2.07** (1.38, 3.09) | 2.10** (1.37, 3.22) |
| *Poor* | 7.18** (4.35, 11.85) | 4.36** (2.30, 8.24) | 6.35** (3.33, 12.09) | 2.86* (1.37, 6.00) | 3.25** (1.92, 5.52) | 3.32** (1.85, 5.94) |
| **Food insecurity** | 1.85** (1.33, 2.58) | 1.70* (1.14, 2.53) | 1.75* (1.10, 2.79) | 1.56 (0.93, 2.61) | 1.61* (1.13, 2.30) | 1.47* (1.01, 2.15) |
| **Family debt** | 2.78** (1.78, 4.34) | 1.68* (1.01, 2.79) | – | – | – | – |
| **Perceived social support** | 0.41** (0.31, 0.54) | 0.61* (0.44, 0.85) | 0.42** (0.29, 0.62) | 0.63* (0.41, 0.96) | – | – |
| *n* for the final model |  | 1028 |  | 1048 |  | 1046 |
| Variance explained |  | 24.2% |  | 21.1% |  | 11.1% |
| Hosmer-Lemeshow test | | $\chi^2 = 994.32$;<br>*p*-value = 0.37 | | $\chi^2 = 830.93$;<br>*p*-value = 0.91 | | $\chi^2 = 392.84$;<br>*p*-value = 0.04 |
| cvMean AUC (95% CI) |  | 0.82 (0.79, 0.85) |  | 0.82 (0.79, 0.86) |  | 0.73 (0.70, 0.77) |

Abbreviations: aOR, adjusted odds ratio; cvMean AUC, cross-validated mean area under the curve for the multivariable model; OR, odds ratio; Ref, reference group
\* *p*-value <0.05, \*\* *p*-value ≤0.001

**Table 4.** Univariate and multivariable analysis of correlates of mental health problems among men, *n=480*.

| Covariate | Positive screen for depressive symptoms |  | Positive screen for anxiety symptoms |  | Positive screen for PTSD |  |
| --- | --- | --- | --- | --- | --- | --- |
|  | Univariate analysis OR (95% CI) | Multivariable analysis aOR (95% CI) | Univariate analysis OR (95% CI) | Multivariable analysis aOR (95% CI) | Univariate analysis OR (95% CI) | Multivariable analysis aOR (95% CI) |
| **Residence** |  |  |  |  |  |  |
| *Rural setting* | – | – | Ref | Ref | – | – |
| *Urban setting* | – | – | 9.47* (1.28, 69.95) | 8.75* (1.17, 65.42) | – | – |
| **Monthly household Income (Ksh)** |  |  |  |  |  |  |
| <10,000 | – | – | – | – | Ref | Ref |
| 10000 – 29999 | – | – | – | – | 0.52* (0.33, 0.84) | 0.49* (0.30, 0.82) |
| >30,000 | – | – | – | – | 0.46 (0.13, 1.59) | 0.40 (0.11, 1.48) |
| **Somatic symptoms** | – | – | – | – | 2.27** (1.44, 3.57) | 1.74* (1.08, 2.83) |
| **Stressful life events** |  |  |  |  |  |  |
| *None* | Ref | Ref | Ref | Ref | Ref | Ref |
| *>5 events* | 1.45 (0.54, 3.91) | 1.47 (0.52, 4.17) | 1.00 (0.22, 4.66) | 1.16 (0.24, 5.48) | 5.58 (0.74, 42.11) | 5.00 (0.66, 38.07) |
| *≥5 events* | 3.15* (1.18, 8.42) | 2.57 (0.90, 7.30) | 2.88 (0.66, 12.69) | 3.65 (0.81, 16.50) | 21.59* (2.90, 160.49) | 17.69* (2.34, 133.46) |
| **Self-perceived wellbeing** | 0.87** (0.83, 0.90) | 0.88** (0.84, 0.92) | – | – | – | – |
| **Sexual abuse** | 3.07** (1.82, 5.18) | 2.29* (1.27, 4.15) | – | – | – | – |
| **Verbal abuse** | – | – | – | – | 2.36** (1.44, 3.86) | 1.71* (1.01, 2.90) |
| **Living in rented houses** | 2.20* (1.29, 3.75) | 2.50* (1.39, 4.48) | – | – | – | – |
| **Days spent on light activities in the previous week** | 1.14* (1.02, 1.27) | 1.16* (1.03, 1.31) | – | – | – | – |
| **Days spent on vigorous activities in the previous week** | – | – | 0.85* (0.73, 0.99) | 0.83* (0.71, 0.98) | – | – |
| **Perceived social support** | 0.46** (0.30, 0.72) | 0.58* (0.36, 0.96) | 0.42** (0.29, 0.62) | 0.39* (0.20, 0.78) |  |  |
| *n* for the final model |  | 480 |  | 480 |  | 480 |
| Variance explained |  | 18% |  | 12% |  | 13.5% |
| Hosmer-Lemeshow test | | $\chi^2 = 341.32$;<br>$p$-value = 0.38 | | $\chi^2 = 26.37$;<br>$p$-value = 1.00 | | $\chi^2 = 26.75$;<br>$p$-value = 0.32 |
| cvMean AUC (95% CI) |  | 0.79 (0.74, 0.83) |  | 0.75 (0.68, 0.83) |  | 0.76 (0.71, 0.81) |

Abbreviations: aOR, adjusted odds ratio; cvMean AUC, cross-validated mean area under the curve for the multivariable model; OR, odds ratio; Ref, reference group
\* $p$ -value <0.05, \*\* $p$ -value ≤0.001

**Table 5.**
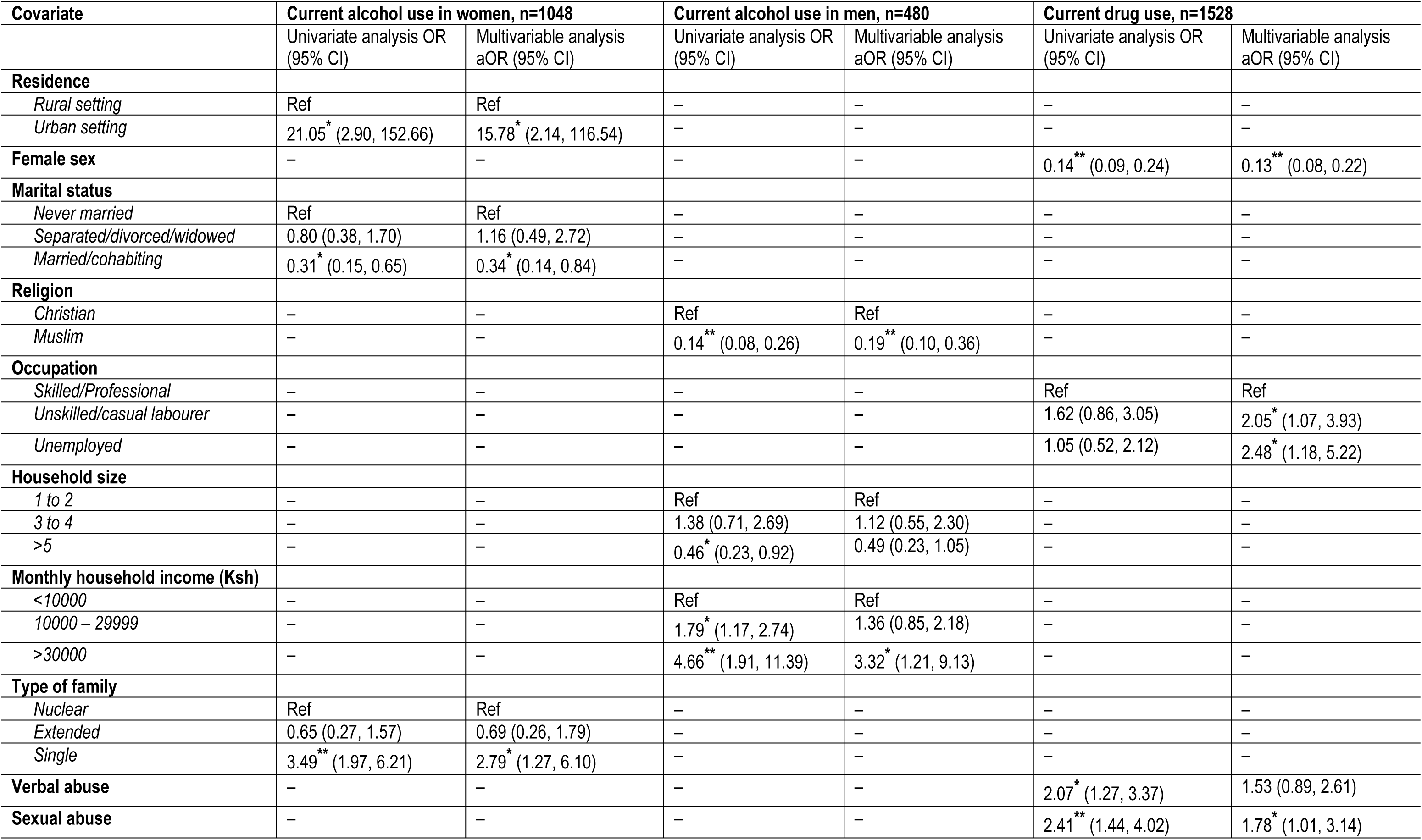
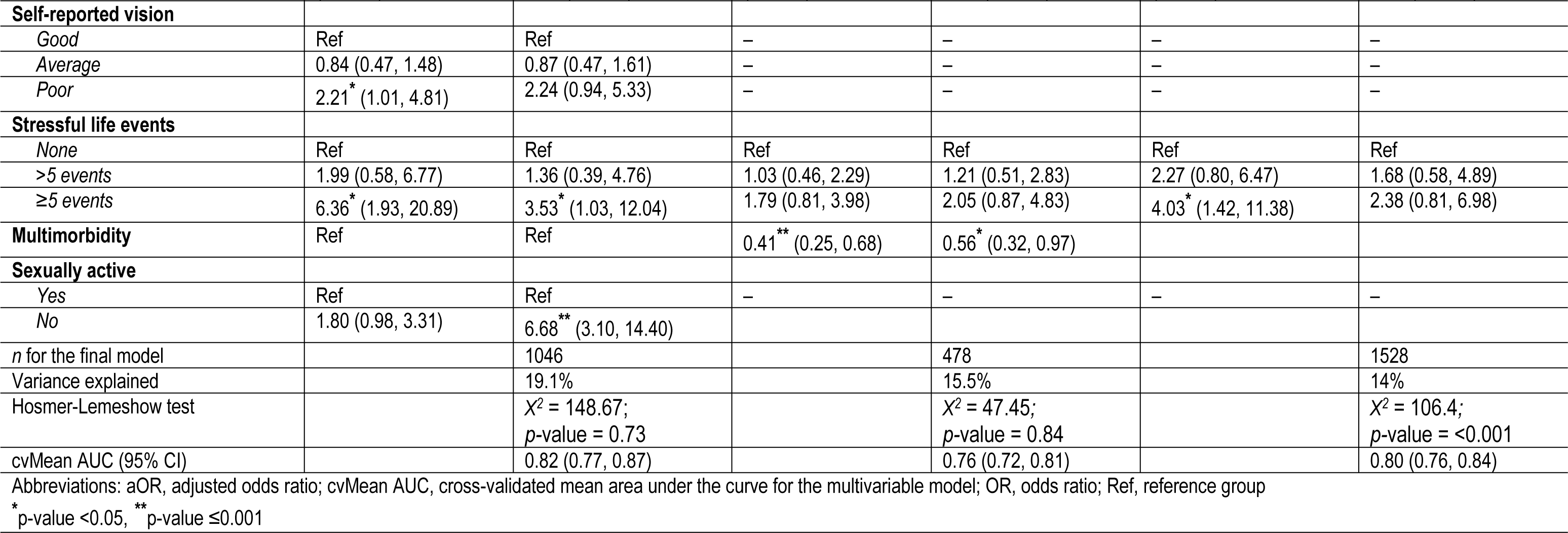
Univariate and multivariable analysis of correlates of current substance use among men and women.

#### Correlates of mental health problems among women

In the final multivariable analysis model (Table 3), urban residence, higher educational status, household food insecurity, household debt, poor self-reported eyesight, experiencing more than five stressful life events, and declining self-rated health status were significantly associated with higher odds of depressive symptoms in women. On the other hand, urban residence, higher educational status, poor self-reported health, experiencing more than five stressful life events, and being unemployed or casual labourer were associated with higher odds of anxiety symptoms among women in the final multivariable model. Increasing self-reported wellbeing and perceived social support were associated with reduced odds of depressive and anxiety symptoms among women. Factors associated with higher odds of PTSD symptoms in the final multivariable analysis included primary/secondary level of education, urban residence, household food insecurity, declining self-rated health status, and experiencing stressful life events. Older age (>50 years) was associated with reduced odds of PTSD symptoms among women.

#### Correlates of mental health problems among men

In the final multivariable analysis (**Table 4**), sexual abuse from a spouse, living in rented houses, and light physical activity were associated with higher odds of depressive symptoms. Conversely, urban residence was associated with increased odds of anxiety symptoms. On the other hand, verbal abuse, somatic complaints, and stressful life events were associated with higher odds of PTSD symptoms among men. Factors associated with reduced odds of mental health problems among men in the final multivariable analysis included increased vigorous exercise, increasing household income, perceived social support, and increasing self-reported wellbeing.

#### Correlates of substance use problems across the sample

In the final multivariable analysis **(Table 5)**, urban residence, experiencing more than five stressful life events, being sexually active, and living in a single-parent family were significantly associated with higher odds of any current alcohol use in women. On the other hand, higher monthly household income was significantly associated with higher odds of any current alcohol use among men. Protective indicators of any current alcohol use included being married/cohabiting (in women), larger household size, being a Muslim, and multimorbidity (among men). Risk indicators for any current drug use across the sample in the final multivariable analysis **(Table 5)** included unemployment/casual work and sexual abuse. Female sex was significantly associated with reduced odds of any current drug use across the sample.

## Discussion

### Key findings

In this study, we documented the prevalence and associated factors of mental and substance use problems in a community sample of adults drawn from Nairobi, Mombasa and Kwale counties in Kenya. Our study is unique in that it encompasses a range of key mental and substance use problems. It gives a comprehensive account of the mental, general health and psychosocial correlates of these conditions. Based on a sample of 1528 adults (69% women), the present study found a relatively high prevalence of depressive symptoms (26%), symptoms of PTSD (21%), anxiety symptoms (11%), and current alcohol use (13%). However, the prevalence of current drug use and past year’s prevalence of hazardous alcohol and drug use were low (<10%) across the sample. Notably, the prevalence of depressive and anxiety symptoms was significantly higher among women compared to their male counterparts. On the other hand, both current and past-year alcohol and drug use problems were significantly higher in men than women. We found no significant differences between women and men in the prevalence of PTSD symptoms. In our second objective, we identified several factors (mostly demographic and psychosocial) that were associated with increased odds of mental and substance use problems, some overlapping between men and women (e.g. urban residence and experiencing stressful life events), others overlapping across conditions (e.g. urban residence, experiencing stressful life events, household food insecurity, and declining self-rated health) while others were unique across sex and conditions. Protective indicators against mental health problems included higher perceived social support, higher subjective wellbeing, older age (>50 years) and higher household income (in both sexes), whereas those of substance use problems included being married/cohabiting (in women) and larger household size, being a Muslim and multimorbidity (among men).

### Prevalence of mental and substance use problems

The prevalence of mental and substance use problems has been shown to vary widely in SSA, partly due to differences in study populations/participants, measures and cut-offs used, as well as study designs [39–43]. Our finding of a 26% prevalence of depressive symptoms is more than the global prevalence of 3.4%, which has been observed for depressive disorders [12]. This estimate is also higher than the prevalence of 4.5%, which has been documented in SSA for depressive disorders [12]. Nonetheless, this figure is within the estimates of 4.2 –50.4%, which has been reported among general populations from several studies in Kenya [18, 44, 45] and similar settings in East Africa [46–50]. The elevated rates of depressive symptoms in our study could be due to several biological, demographic and psychosocial vulnerabilities, some of which have been confirmed through this study. Generally, fewer studies have examined the prevalence of anxiety and PTSD relative to other CMDs in Kenya and the wider SSA region. Our study directly addresses this gap in Kenya and the region. Likewise, the reported prevalence of anxiety in our study (11%) is more than the global prevalence (4%) [51]. It is also worth noting that the reported prevalence in this study is lower than that reported in a previous study (20%), which was conducted in a similar setting but during the COVID-19 pandemic in Kenya [44]. On the other hand, the reported prevalence of PTSD symptoms in this study (21%) is more than the lifetime prevalence of 3.9%, which has been documented by the World Mental Health Surveys [52] but comparable to the prevalence of 22% reported in a systematic review and meta-analysis of studies conducted in SSA [42]. The observed prevalence is also higher than previously reported estimates in Kenya: 11% in a cross-sectional household survey in 2015 [53] and 16% among university students in 2015 [54]. However, our prevalence is lower compared to some previous reports in Kenya: 65% among healthcare providers in the early phase of COVID-19 [55], 62% among internally displaced persons [56] and 65% among patients [57]. This is expected given that the previous studies assessed high-risk populations. Our observations also confirm previous findings that the prevalence estimates of probable PTSD are highly variable across settings partly because of differences in study populations, contexts/regions, study designs, etc., thus highlighting the need for context-specific studies.

The prevalence estimates for current substance use observed in this study are lower than what has been previously reported in the literature in Kenya [58–60]; however, our estimates (for alcohol use) are comparable to the latest national estimates as per the national survey on the status of drugs and substance use in Kenya (NACADA) reports [61]. Similarly, the observed past-year prevalence estimates of hazardous drug use are lower than what has been reported previously in Kenya [61]. Variations in study populations (e.g. age), source of information (self-report versus other formats), study setting, and assessment instruments could partly explain the variances observed in the literature. The observations could also be due to the community sensitization on drugs and alcohol across the counties.

In our sex-stratified analysis, women presented with significantly higher prevalence estimates for depressive and anxiety symptoms than men; however, men presented with significantly higher levels of both current and past-year prevalence of alcohol and drug use problems. Our observation is consistent with previous findings in the literature [62]. These differences can partly be accounted for by biological and psychosocial vulnerabilities in the respective genders. Despite the observed differences in depressive and anxiety symptoms between men and women in this study, our study points to a narrowing in the sex-stratified prevalence of these problems in this setting – emphasizing the importance of addressing these conditions in both men and women. Few studies have aggregated their results by sex in Kenya; hence, more studies will provide more insights into this finding.

### Correlates of mental and substance use problems

Assessing sex disparity in mental health research is crucial for targeted support and intervention, yet often overlooked in Kenya and other settings in SSA. In addressing this gap, we stratified our analysis of the correlates of mental and substance use problems by sex. Urban residence and experiencing stressful life events were the only risk indicators common to both men and women for mental and substance use problems. Generally, the influence of urban environments on mental health is not well understood. There is some evidence showing that urban residence is associated with higher risks of mental health problems – especially common mental disorders and substance use disorders [63], although results are contradicting [64]. The finding that experiencing stressful life events was associated with higher odds of mental and substance use problems is supported by results from previous research across different settings [65–67]. Our findings also revealed an overlap of some of the risk indicators (e.g. urban residence, stressful live events, food insecurity and declining self-rated health) across mental and substance use problems. This potentially suggests that individuals who experience these shared cumulative risk indicators are more likely to face multiple mental health challenges. Furthermore, it also implies that actions taken to address any of these shared risk factors are likely to have a promotive spillover effect across the different mental health problems.

Risk indicators unique to women’s mental health problems included household debt, poor self-reported eyesight, being unemployed or casual labourer, and higher educational status (relative to none). On the other hand, being sexually active and single-parent households were uniquely associated with higher odds of current alcohol use in women. These findings are supported by previous research in different contexts [68–76]. Among men, the unique risk indicators of mental health problems included sexual and verbal abuse from a spouse, living in rented houses, somatic complaints and taking part in light physical activity. Higher monthly household income was significantly associated with higher odds of current alcohol use in men. The finding that sexual and verbal abuse was a significant risk indicator for mental health problems in men but not women was a surprise finding in this study. Emerging data from empirical research and anecdotal reports shows that men in SSA are experiencing increasing ’hidden’ reports of sexual and other forms of violence [77, 78]. In the present study, men reported slightly higher frequencies of both sexual abuse (16% vs 13%) and verbal abuse (58% vs 53%) incidences than women (though not statistically significant). We propose further studies to explore this finding. The association of somatic complaints and living in rented houses with higher odds of mental health problems is consistent with previous research findings [79–81]. The association between light-intensity physical activity and mental health problems is generally mixed in literature [82]. Additional studies are needed to understand the underlying mechanisms of this relationship.

Given the low prevalence estimates of drug use problems – we did not disaggregate the sample by sex when investigating the correlates of drug use in the study. Identified risk indicators for any current drug use included unemployment/casual work and sexual abuse – all of which are consistent with previous research [83, 84]. A follow-up study with a larger sample size is encouraged to fully explore the determinants of drug use problems in this setting.

Identifying protective factors for mental and substance use problems is essential for the development of interventions/programs designed to prevent or lower the burden that these conditions place on individuals and provide future directions for public health policy. Higher perceived social support, older age (>50 years), increasing self-reported wellbeing, higher household income and vigorous physical activity were significantly associated with reduced odds of mental health problems. On the other hand, being married/cohabiting, larger household size, being a Muslim and having multimorbidity were significantly associated with reduced odds of current alcohol use. Our findings are consistent with prior research.

### Limitations

Our findings should be interpreted in light of certain limitations. We utilised screening measures to assess the prevalence and correlates of mental and substance use problems. Previous research has shown that screening tools have the potential of over-reporting the burden of disorders partly because of using unvalidated instruments. To minimize this, all the measures used have been adequately adapted and/or validated in similar settings in Kenya. Relatedly, we did not use audio-computer-assisted self-interview (ACASI) when assessing substance use problems in the study, and this may have introduced social desirability bias when participants were responding to the substance use questions. Moreover, the tool we utilised to assess drug use problems (DUDIT) does not highlight individual drug types; thus, we could not provide individual drug use prevalence estimates. We recommend a follow-up study to provide clarifications on this account. Additionally, given the low prevalence estimates of drug use, we could not offer sex-stratified correlates of drug use in this sample – future studies should utilize larger sample sizes to provide clarity on this. Also, the cross-sectional design of the study limits any inferences on causality in the observed associations.

### Implications

Despite the limitations above, the findings of this study have crucial implications for mental health policy, practice and further research relating to adults living in rural and urban informal settlements. The World Health Organization has called for countries to fill gaps in knowledge about mental health, and the present study addresses this call. Our study found high prevalence estimates of common mental disorders and significant levels of substance use problems in this sample (with distinct patterns across sex), which was partly driven by multiple overlapping and unique risk indicators. Though we did not assess access to mental health services, it is likely that there was minimal, if any, access to and utilization of these services by the participants, given the high mental health treatment gap (75%) currently being reported in Kenya [18]. These findings emphasize the urgent need for preventive, promotive and curative mental health services to address the existing mental health burden – this will, however, need significant funds to support these efforts [85]. Low-intensity lay-administered psychological interventions such as Problem Management Plus intervention, which have shown promise in feasibility studies in this setting, can be scaled up to maximize their reach among the population [86, 87], given the huge gap in trained mental health specialists. A task-shifting framework for the delivery of comprehensive, collaborative, and community-based mental health care in LMICs has been proposed to expand mental health services [88]. Pathways of proper and timely management of these problems are also very important in ensuring continuity of care. The Kenya National Mental Health Plan 2021-2025 provides a framework for the national and county governments to begin taking steps in timely assessment and management, including preventive and promotive mental health care, for different populations in the country. Our study also points out the significant disparities in the prevalence and correlates of mental and substance use problems in this setting, thus emphasizing the need for tailored and gender-sensitive mental health strategies to address the unique issues faced by men and women in this setting.

Our study identified several factors that were significantly associated with mental and substance use problems among the participants. The majority of the factors were psychosocial and potentially amenable with appropriate psychosocial interventions [89, 90], and they can also be useful targets in preventive and promotive mental health strategies. Given the cross-sectional design of our study, our findings on risk and protective indicators preclude any conclusions on causality. Further studies with appropriate designs that can allow causal inferences are needed to corroborate our findings.

## Conclusions

Understanding the gender-stratified prevalence and correlates of mental and substance use problems is relevant in guiding researchers, policymakers and practitioners to adequately put in place gender-specific interventions. The current study highlights important gender disparities in the burden and determinants of mental and substance use problems among adults in rural and urban informal settlements in Kenya. Women presented with significantly higher levels of depressive and anxiety symptoms than their male counterparts, although the magnitude was smaller. On the other hand, men presented with significantly higher levels of both current and past-year alcohol and substance use compared to women. We also identified several risk and protective indicators for these conditions, the majority of which were demographic and psychosocial factors. Some of these factors overlapped across sex and studied conditions, while others were unique. These results underscore the urgent need for tailored and gender-distinct mental health strategies in addressing these issues in the study population and also the need to address underlying societal and structural issues that drive the observed gender disparities.

## Acknowledgements

We are grateful to the study participants who took part in this study. We would also like to thank all the field enumerators including Joseph Muye, Eunice Kisau, Joyce Jefwa, Sarah Minoo, Juliet Adek, Peter Wanyama, Emmanuel Kombo, Joel Owaret, Emmah Joy, Mercy Juliet, Peter Mulupi, Mercy Cherotich, John Maina, Kevin Wekesa, Felix Kipkosgei, Elly Opar, Isaac Lihanda, Faith Kwamboka, Tom Joe and Ann Muisyo for their role in data collection. We also appreciate all the study mobilizers including the community health volunteers, community health assistants, community focal persons from the study collection sites.

## Data Availability

The de-identified dataset used and analysed during this current study will be made available upon reasonable request, in due consideration of Aga Khan University’s data sharing policies. Requests to access the datasets should be directed to.

## Author contributions

AA conceptualized and designed the study. AA and PNM developed study protocols, tools, and training programmes. RO programmed the study questions on tablets and managed study data throughout the data collection period. PNM conducted training on the enumerators and coordinated the data collection process. AK and MM supervised data collection in Nairobi County. PNM and AK conducted data cleaning and analysis. PNM wrote the first draft of the manuscript. All authors critically reviewed subsequent versions of the manuscript and approved the final version for submission. All authors read and approved the final manuscript.

